# Circulating prothymosin alpha and immunoglobulin G3 in acute rheumatic fever and rheumatic heart disease: A case-control study

**DOI:** 10.1101/2025.06.24.25330185

**Authors:** Gul Afshan, Nicole Tsui, Humera Javed, Tehmina Kazmi, Emma Ndagire, Jafes Pulle, Kristin Huse, Amelia Lias, Shagorika Talukder, Natalie Lorenz, Reuben McGregor, Craig Sable, Andrea Z. Beaton, Nicole J. Moreland, Emmy Okello, Masood Sadiq, Tom Parks

## Abstract

The lack of specific diagnostic tests for acute rheumatic fever (ARF) and its consequence rheumatic heart disease (RHD) is a key barrier to effective control of these diseases in low resource settings. Prothymosin alpha (PTA) and immunoglobulin G3 (IgG3) have recently been proposed as ARF biomarkers, although studies of these markers have so far been limited to single populations. Consequently, we evaluated PTA and IgG3 in patients with ARF, RHD and controls from studies set in Pakistan and Uganda.

## Research Letter

Acute rheumatic fever (ARF) and its chronic consequence rheumatic heart disease (RHD) are important causes of cardiovascular death and disability across much of the world (1). Control of these diseases is complicated by the uncertain performance of the Jones’ Criteria (2) in low resource settings, exacerbated by an absence of specific diagnostic tests for ARF. Prothymosin alpha (PTA) was recently proposed as a key contributor to RHD immunopathogenesis, as well as a putative biomarker discriminating patients with chronic RHD from healthy controls (3). Therefore we sought to assess whether blood PTA was elevated in patients with ARF or RHD compared to unwell controls, indicating potential as a diagnostic. We compared this to immunoglobulin G3 (IgG3), which was recently shown to be elevated in ARF, as an additional putative biomarker (4).

Our study was approved by institutional review boards at the Children’s Hospital Lahore, Pakistan, and Makerere University, Kampala, Uganda, and the Imperial College London Research Ethics Committee. We undertook a case-control study investigating genetic susceptibility to ARF and RHD at the Children’s Hospital Lahore between August 2019 and March 2022, with an interruption in March 2020 due to COVID-19. Here we focus on serum samples from children and young adults aged 5-21 years attending hospital with ARF, RHD or non-ARF diagnoses obtained from the study after September 2020.

For this investigation, we selected 30 individuals with suspected ARF comprising all 16 in the cohort with C-reactive protein (CRP) greater than 1mg/dL and an additional 14 with CRP below this threshold. For comparison, we selected all 12 control individuals with non-ARF diagnoses and CRP greater than 1mg/dL, and 14 non- inflammatory control individuals with CRP less than 1mg/dL matched where possible by age and sex to the inflammatory cases. Finally, we included all four available samples from children with RHD without concurrent ARF giving a total sample of 60 individuals.

We measured PTA in duplicate in undiluted sera using a sandwich ELISA kit according to the manufacturer’s instructions (LSBio; Cat. F50952). Total IgG3 was measured in sera diluted up to 1:50,000 using a bead assay according to the manufacturer’s instructions (Merck; Cat. HGAMMAG-301K) (5). Preceding streptococcal infections were defined based on an anti-streptolysin O titer of greater than 250 IU/ml measured by automated immunoassay (Abbott, USA). Neither group A streptococcal rapid antigen tests nor anti-DNase B titer measurement were available to the study. The mean rank of groups was compared using a Kruskal- Wallis test while specific groups were compared using the Wilcoxon rank-sum test and multivariate analyses were performed by logistic regression in R software (version 4.4). Sample size was determined by the number of samples available at the time of the study rather than a formal power calculation.

The median age was 12 years and 37 of 60 were male (Table 1). All 30 suspected ARF cases had evidence of carditis, 22 had arthritis or polyarthralgia, and none had chorea. For analysis, we reclassified the 30 suspected ARF cases using the 2015 Jones’ Criteria (2), giving 15 definite cases (all recurrent) and 15 possible (11 recurrent) not completely fulfilling the diagnostic criteria. A single individual was excluded due to an invalid IgG3 measurement but otherwise there were no missing data. Total IgG3 was significantly elevated in the definite ARF cases compared to both inflammatory (p=0.006) and non-inflammatory controls (p=0.015; Figure 1a); however, PTA concentrations were similar across all five groups (p=0.56; Figure 1b), irrespective of the presence or absence of an inflammatory state.

**Table 1.** Characteristics of the Pakistan cohort.

|  | Non-inflammatory control (n=9) | Inflammatory control (n=17) | Possible ARF (n=15) | Definite ARF (n=15) | Chronic RHD (n=4) |
| --- | --- | --- | --- | --- | --- |
| Age (years), median (IQR) | 13 (11-17) | 10 (9-14) | 12 (10-13) | 12 (10-14.5) | 10.5 (10-11) |
| Female, n (%) | 4 (44) | 8 (47) | 6 (40) | 3 (20) | 2 (50) |
| Carditis, n (%) | 0 | 0 | 15 (100) | 15 (100) | * |
| Joint involvement, n (%) | † | † | 9 (60) | 13 (87) | 0 |
| CRP (mg/L), median (IQR) | 3 (0-7) | 30 (3-47) | 3 (1-13) | 20 (10-42) | 0 (0-1) |
| ASO titer, median (IQR) | 152 (104-199) | 127 (67-215) | 150 (53-214) | 477 (311-601) | 80 (61-119) |

\*Chronic RHD without acute carditis
†Details of joint symptoms not recorded

**Figure 1:**
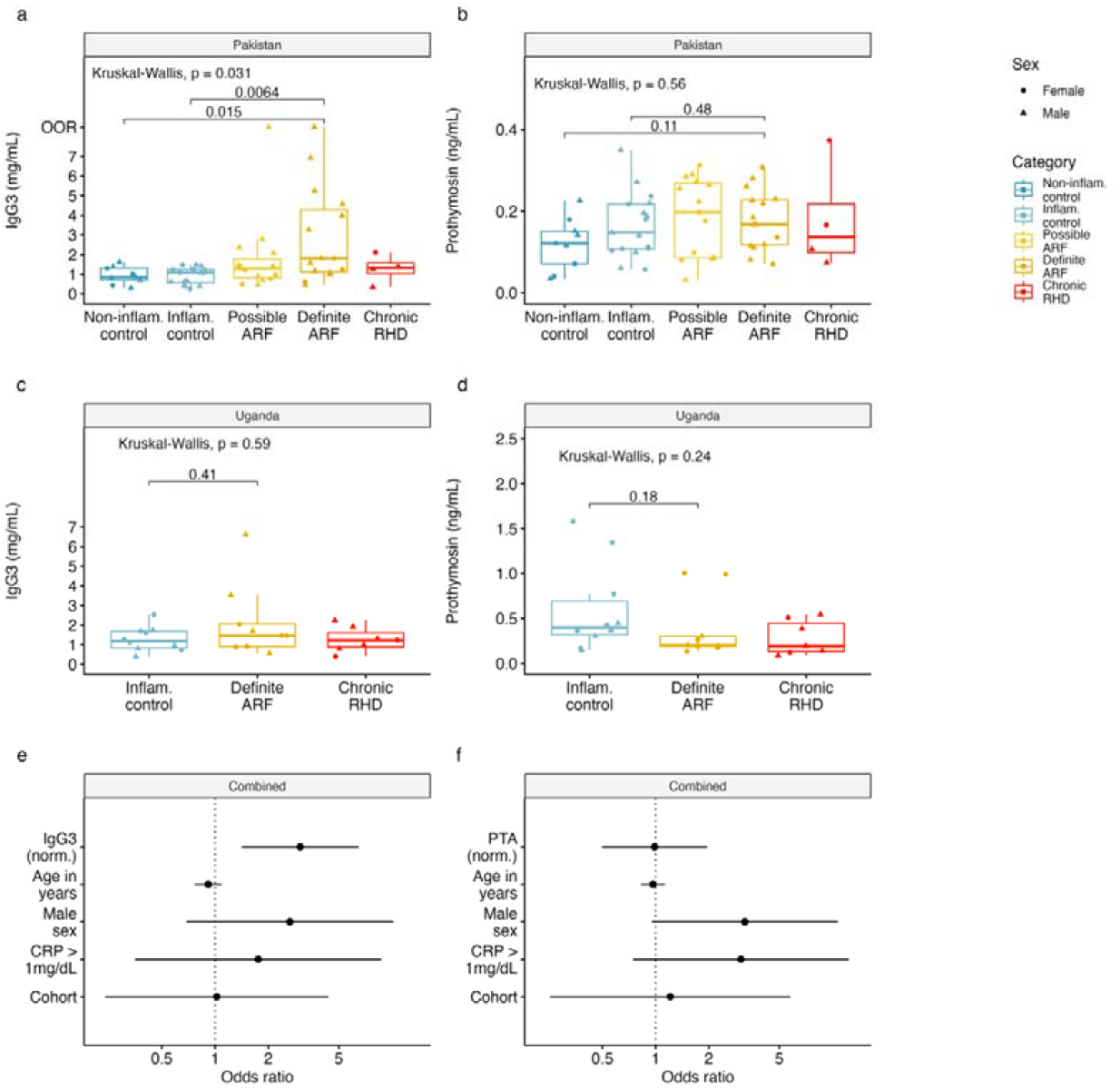
Elevation of IgG3 but not PTA in ARF. Distribution across study groups of: **(a)** IgG3 in serum samples from Pakistan; **(b)** PTA in serum samples from Pakistan; **(c)** IgG3 in plasma samples from Uganda; **(d)** PTA in plasma samples from Uganda. Multivariate logistic regression model based on both cohorts comparing definite ARF (n=25) to the inflammatory and non-inflammatory controls group combined (n=36). Odds ratios and 95% confidence are shown for each one unit increase in: **(e)** normalized IgG3; **(f)** normalised PTA. Normalization was performed using a Yeo-Johnson Transformation and models are adjustment was made for age in years, male sex, CRP greater than 1mg/dL and the cohort. OOR, beyond upper limit of detection

The previous study detected PTA in plasma rather than serum (3). Accordingly, to confirm the lack of elevation of PTA in the Pakistan cohort was not due our use of sera, we obtained plasma samples from a previously reported prospective epidemiological study of ARF in two regions of Uganda. That study ran between January 2018 and February 2020, recruiting children aged 3-17 years with suspected ARF (6). For our analysis, we measured PTA in duplicate in plasma diluted 1:4 and IgG3 in serum diluted 1:50,000 as described above. Across the cohort, the median age was eight years and half were male (Table 2). We included nine individuals with definite ARF (eight with CRP greater than 1mg/dL), 10 controls with known or unknown non-ARF diagnoses (all CRP greater than 1mg/dL), and seven individuals with chronic RHD. Six of the definite ARF cases had carditis while the remaining three had arthritis or polyarthralgia.

**Table 2.** Characteristics of the Uganda cohort.

|  | Inflammatory control (n=10) | Definite ARF (n=9) | Chronic RHD (n=7) |
| --- | --- | --- | --- |
| Age (years), median (IQR) | 7 (5-10) | 7 (6-10) | 10 (7.5-13) |
| Female, n (%) | 6 (60) | 5 (56) | 2 (29) |
| Carditis, n (%) | 0 | 6 (33) | * |
| Joint involvement, n (%) | 9 (90) | 7 (78) | 1 (14) |
| CRP (mg/L), median (IQR) | 44 (29-46) | 53 (37-85) | 2 (2-15) |
| ASO titer, median (IQR) | 115 (81-203) | 369 (221-676) | 188 (145-315) |

\*Chronic RHD without acute carditis

Overall plasma PTA concentrations in the samples from Uganda (median 0.3ng/mL, IQR 0.18-0.49) were higher than in the serum samples from Pakistan (median 0.17ng/mL, IQR 0.1-0.22); however, plasma PTA concentrations were similar across groups within the Ugandan study (p=0.24; Figure 1c), as well as in a combined analysis of the two cohorts (p=0.52). While there was no significant difference in median serum IgG3 across groups within the Ugandan study (p=0.59; Figure 1d), the median IgG3 serum concentration was significantly higher among definite ARF cases compared to controls in a combined analysis (1.63mg/mL vs 1.07mg/mL, p=0.001). Additionally, the relationship between ARF and IgG3 was robust to adjustment for age, sex, cohort and inflammatory status (p=0.004; Figure 1e), while no such relationship was apparent for PTA (p=0.94; Figure 1f).

In summary, we found blood PTA levels to be similar across ARF, RHD and controls, conflicting with a previous report that found PTA was elevated in RHD compared to healthy controls (3). In contrast, total IgG3 was elevated in ARF compared to controls, independent of the acute phase response. To our knowledge, this is the first time elevation of IgG3 in ARF has been described outside of New Zealand, although the difference we observed here was less marked than that reported in previous studies (4,5). While our relatively small sample size was a limitation, it was sufficient to replicate the previously observed difference for PTA, and our study benefited from samples from two distinct ARF studies, inclusion of a spectrum of ARF in addition to RHD, a comparison with unwell controls, and use of both serum and plasma.

It remains unclear why our findings relating to PTA differ from those in the previous report (3). However, importantly, we used an updated version of the ELISA kit produced by the same manufacturer, designed to detect PTA at 10-fold lower concentrations than that used in the previous study. Although we have not compared the performance of the two products, it is notable that the version that we used was based on mouse monoclonal rather than rabbit polyclonal capture antibodies. Nonetheless, we only had sera available from Pakistan, while our analyses based on plasma were limited to the smaller Ugandan cohort. Accordingly, it is possible we had insufficient power to detect a difference that was specific to plasma, although it is noteworthy that patients with both definite ARF and chronic RHD from Uganda had lower median concentration of PTA than controls, rendering it unlikely that PTA could form the basis of a diagnostic test.

A total of 19 of 24 (79%) children with definite ARF from Pakistan and Uganda had IgG3 above the previously highlighted threshold of 1mg/ml. While this is lower than the proportion reported in the study from New Zealand (4), there are differences in the clinical characteristics of the cohorts recruited in Uganda, Pakistan and New Zealand, which may explain this discrepancy. Importantly, we used the definition of definite ARF for moderate and high-risk populations from the latest Jones Criteria revision, which included adjustments intended to improve sensitivity (2). The impact of these criteria was especially apparent in Uganda since four of the nine children in the definite ARF category had polyarthralgia as a standalone major manifestation. It is notable that a higher proportion of children who met the stricter criteria for ARF intended for low-risk populations – 16 of 19 (84%) – had IgG3 above the 1mg/ml threshold, approaching the proportion reported from New Zealand (4). Although IgG3 is not sufficiently elevated in our analysis to serve as a standalone biomarker, the consistency of our findings from two distinct cohorts with the previous studies from New Zealand is striking, and suggests further work is justified to understand the relationship between the disease and this subclass of antibody.

Accordingly, there is a clear need for ongoing work to identify biomarkers for ARF, including measurement of IgG3 and other biomarker candidates in additional cohorts, which we are now undertaking through the Acute Rheumatic Fever Diagnostic Network (arcdiagnosticnetwork.org).

## Acknowledgements

We thank the patients and families who participated in the study. We also thank the Acute Rheumatic Fever Diagnostic Network.

## Contributions

TP is the guarantor and accepts full responsibility for the work. TP, MS, EO, NJM, AB, CS and RM conceived the study. GA, NT, HJ, TK, EN, JP, KH, AL, ST and NL acquired the data. GA, NT and TP collated and analysed the data. TP, GA, NT, MS, EO, NJM and AB interpreted the findings. TP wrote the first draft of the manuscript. All authors contributed to and critically revised the manuscript. All authors approved the final version.

## Data Availability

The data that support the findings of this study are available from the corresponding authors upon reasonable request.

## Note

This research was funded in part by the Wellcome Trust [222098/Z/20/Z]. For the purpose of open access, the author has applied a CC BY public copyright licence to any Author Accepted Manuscript version arising from this submission.

